# Insulinotropic effect of endogenous incretins is greater after gastric bypass than sleeve gastrectomy despite diminished beta-cell sensitivity to plasma incretins

**DOI:** 10.1101/2023.03.28.23287755

**Authors:** Marzieh Salehi, Richard Peterson, Devjit Tripathy, Samantha Pezzica, Ralph DeFronzo, Amalia Gastaldelli

**Affiliations:** Division of Diabetes, University of Texas at San Antonio, San Antonio, TX, United States; STVHCS, Audie Murphy Hospital, San Antonio, TX, United States; Department of Surgery, University of Texas at San Antonio, San Antonio, TX, United States; Cardiometabolic Risk Unit, CNR Institute of Clinical Physiology, Pisa, Italy

**Keywords:** Gastric bypass, sleeve gastrectomy, β-cell function, incretin, endogenous GLP-1, hypoglycemia

## Abstract

**Background/Aims:** Prandial hyperinsulinemia after Roux-en Y gastric bypass surgery (GB), and to lesser degree after sleeve gastrectomy (SG), has been attributed to rapid glucose flux from the gut and increased insulinotropic gut hormones. However, β-cell sensitivity to *exogenous* incretin is markedly reduced after GB. This study examines the effect of GB versus SG on prandial glycemia and β-cell response to increasing concentrations of *endogenous* incretins.

**Methods:** Glucose kinetics, insulin secretion rate (ISR), and incretin responses to 50-gram oral glucose ingestion were compared between 10 non-diabetic subjects with GB versus 9 matched individuals with SG and 7 non-operated normal glucose tolerant controls (CN) on two days with and without administration of 200 mg sitagliptin.

**Results:** Fasting glucose and hormonal levels were similar among 3 groups. Increasing plasma concentrations of endogenous incretins by 2-3-fold diminished post-OGTT glycemia and increased β-cell secretion in all 3 groups (p<0.05), but insulin secretion per insulin sensitivity (i.e., disposition index) was increased only in GB (p<0.05 for interaction). As a result, sitagliptin administration led to hypoglycemia in 3 of 10 GB. Yet, plot of the slope of ISR versus the increase in endogenous incretin concentration was smaller after GB compared to both SG and CN.

**Conclusion:** Augmented glycemic-induced β-cell response caused by enhanced incretin activity is unique to GB and not shared with SG. However, the β-cell sensitivity to increasing concentrations of *endogenous* incretin is smaller after bariatric surgery, particularly after GB, compared to non-operated controls, indicating a long-term adaptation of gut-pancreas axis after these procedures.

**HIGHLIGHTS:** *What is known?:* Glycemic effects of gastric bypass (GB) and sleeve gastrectomy (SG) is attributed to rapid nutrient flux and enhanced insulinotropic effects of gut hormones but β-cell sensitivity to *exogenous* GLP-1 or GIP is diminished after GB.

*What the present findings add?:* Post-OGTT β-cell sensitivity to enhanced *endogenous* incretins by DPP4i is markedly reduced in bariatric subjects versus non-operated controls, and yet insulin secretory response (disposition index) is increased leading to hypoglycemia in GB and not SG.

*Significance?:* Blunted sensitivity to GLP-1 may represent β-cell adaptation to massive elevation in GLP-1 secretion following bariatric surgery to protect against hypoglycemia. The differential effect of enhanced concentrations of incretins on post-OGTT insulin response (disposition index) among GB versus SG highlights a distinct adaptive process among the two procedures. Augmented insulinotropic effects of gut hormones on postprandial insulin secretory response after GB despite a reduced beta-cell sensitivity to plasma concentrations of GLP-1 makes a case for non-hormonal mechanisms of GLP-1 action after GB. Better understanding of long-term effects of bariatric surgery on gut-pancreas axis activity is critical in development of GLP-1-based strategies to address glucose abnormalities (both hyperglycemia and hypoglycemia) in these settings.

## INTRODUCTION

The two bariatric procedures, Roux-en Y gastric bypass (GB) and sleeve gastrectomy (SG) have been widely used for the treatment of diabetes in obese patients. Resolution of T2D following these surgeries is attributed to the amount of weight loss(1, 2), mainly by improving fasting insulin kinetics(3). However, GB, and to lesser extent SG, leads to improved glucose tolerance shortly after surgery (4, 5, 6, 7). Improved glucose tolerance after GB has been attributed to enhanced in nutrient transit resulting in a shift of glucose fluxes, with resetting of the balance between glucose appearance and clearance(2, 8, 9, 10). Emptying of the stomach pouch in individuals after GB is ∼25 times faster than that in non-surgical healthy controls(11). SG reduces the half-time of gastric emptying for both liquid and solid food(12) compared to non-surgical healthy controls (CN), but this effect is smaller than what has been shown after GB. Faster nutrient emptying after bariatric surgery is associated with earlier and higher peaks of glucose and lower nadir glucose levels in parallel with earlier and larger insulin and glucagon-like peptide 1 (GLP-1) response(2, 4, 8, 9, 10, 13, 14, 15). The most extreme effect of GB on glucose metabolism is exemplified by the syndrome of hyperinsulinemia hypoglycemia(10, 16, 17).

Postprandial hyperinsulinemia in GB has been attributed to the combined effects of elevated glucose (8, 10) and a larger incretin effect(5, 13, 18) while the role of the incretin effect after SG is less characterized. Previously, using a GLP-1 receptor (GLP-1R) antagonist, we demonstrated that the contribution of endogenous GLP-1 to postprandial β-cell secretion during a hyperglycemic clamp is 2- to 3-fold larger in GB than matched non-operated controls(18). However, blocking GLP-1R during meal ingestion in GB patients had only a modest effect on prandial insulin secretion rate (ISR) compared to non-operated controls despite a 10-fold increase in GLP-1 secretion (10, 19, 20). Because of this discrepancy, we examined the effect of infusion of increasing doses of GLP-1 or GIP on glucose-stimulated ISR and found that in non-diabetic GB subjects the beta-cell sensitivity to *exogenous* incretins is markedly reduced(21). However, the relationship between incretin secretion and incretin sensitivity within each subject could not be examined, nor did this study address the effect of GB on β-cell effects of increasing *endogenous* incretins (*active*) in this population.

The current study was designed to determine the role of enhancing *endogenous* incretin secretion on β-cell function and glucose fluxes following GB and the differential effect of GB versus SG on endogenous incretin-stimulated ISR in the fed state. We used 200 mg of sitagliptin, which is shown to induce a near-complete inhibition of dipeptidyl peptidase-4 (DPP4) activity within 2 to 12 h post-dose(22), and examined the acute effect of DPP4 inhibitor (DPP4i) on post-OGTT glucose fluxes and islet-cell (insulin and glucagon) and incretin hormonal secretion in three groups of non-diabetic subjects with history of GB, SG, and non-operated controls.

## RESEARCH DESIGN AND METHODS

### Subjects

Ten subjects with previous GB and 9 BMI- and age-matched individuals with SG as well as 7 healthy controls without prior gastrointestinal surgery (CN) were consecutively recruited based on their response to our enrollment effort. All subjects were free of diabetes or renal dysfunction or liver disorder, and none took any medications that interfere with glucose metabolism. The control subjects had no personal or family history of diabetes and had a normal oral glucose tolerance test. Subjects were weight stable for at least 3 months prior to enrollment. The Institutional Review Board of the University of Texas Health at San Antonio approved the protocol (HSC20180070H) and all participants provided written informed consent before participation.

### Experimental protocols

Studies were performed at the Bartter Clinical Research Unit at Audie Murphy VA Hospital in the morning after an overnight fast. Participants were instructed to maintain normal carbohydrate ingestion for 3 days before each visit, and refrain from excessive physical activity. Body composition was assessed using dual-energy X-ray absorptiometry, and waist circumference was measured. Intravenous catheters were placed in each forearm vein for the blood withdrawal and the infusion of glucose tracer. The forearm used for blood sampling was continuously warmed with a heating pad. Throughout the studies blood samples were collected at timed intervals and stored on ice; plasma was separated within 60 min and stored at -80º C until assayed.

After an overnight fasting, fasting blood samples were drawn. At -120 min, subjects received either a single dose of sitagliptin 200 mg orally or nothing (control study), and a primed-continuous infusion of [6,6-^2^H_2_]glucose (28 μmol/kg prime and 0.28 μmol/kg/min constant) was initiated and continued for the remainder of the study(10). At 0 min, a 50 g oral glucose solution mixed with 1 g of uniformly-labeled glucose [U-^13^C]glucose and 1 g of dissolved acetaminophen was consumed within 10 min. These studies were performed in a random fashion, so that half of the subjects received sitagliptin first. Heart rate (HR) and systolic and diastolic blood pressure (SBP and DBP) were monitored throughout the studies and values were averaged over 5-15 min intervals.

### Assays

Blood samples were collected in EDTA tubes for measurement of insulin, acetaminophen, glucose and in aprotinin/heparin/EDTA for assay of C-peptide, glucagon, GLP-1, and GIP and diprotin A for the measurement of *active* GLP-1(23). Plasma glucose was determined using Analox GM9 Glucose Analyzer (Analox Instruments, Stourbridge, UK). Insulin (DIAsource, Neuve, Belgium), C-peptide and glucagon (Millipore, Billerica, MA) were measured by commercial radioimmunoassay. *Active* GLP-1 and total GIP were measured using commercial Multiplex ELISA (Millipore, Billerica, MS), and total GLP-1 using ELISA (Mercodia, Uppsala, Sweden) according to the manufacturers’ specifications. Tracer enrichment was measured by GC-MS (5975, Agilent, Santa Clara, CA) as previously described (10, 24, 25) and acetaminophen was measured by GC–MS using Acetaminophen (acetyl-^13^C_2_,^15^N) as internal standard (Cambridge Isotope Laboratories, Boston, USA) utilizing the same derivatization method used for glucose tracers (26) and monitoring peak of mass 151-154.

### Calculations and analyses

Fasting plasma glucose and hormone concentrations represent the mean of 3 samples drawn before administration of sitagliptin, and the pre-meal values represent the average of the samples drawn over the 10 min before the test meal. Insulin secretion rates (ISRs) were calculated from C-peptide concentrations using deconvolution with population estimates of plasma C-peptide(27). Beta-cell glucose sensitivity (BGS) was calculated in each subject as the slope of ISR and blood glucose concentration for the first part of the OGTT, as glycemia rose to peak value, and for the latter part of the OGTT as glucose concentration declined towards fasting level.

Rates of total glucose appearance (T*Ra*), glucose disappearance (*Rd*), systemic appearance of ingested glucose (*RaO*), and endogenous glucose production (*EGP*) were derived from plasma [6,6-^2^H_2_]glucose and [U-^13^C]glucose enrichments as previously described using the Steele equation(10, 24) and cumulative values for 1 and 3 hours after meal ingestion were estimated. Metabolic clearance of glucose (*MCR*_*Glucose*_) was measured as *Rd*/plasma glucose(28).

Using the trapezoidal rule, the post-OGTT incremental area under the concentration curve (AUC) of islet-cell and GI hormones was calculated from 0-180 and 0-60 minutes to evaluate the total and early responses, respectively, given the altered prandial response pattern after bariatric surgery. Fasting insulin sensitivity was calculated by HOMA-IR(10), pre- and post-OGTT insulin sensitivity were calculated as the ratio of premeal *MCR*_*Glucos*e_/insulin and the prandial AUC of the *MCR*_*Glucos*e_/insulin, respectively(28). Insulin extraction and clearance rates were calculated as previously described (27). Disposition index was calculated as the product of AUC ISR and *MCR*_*Glucose*_/insulin during the 3 hours after oral glucose ingestion(26, 28).

#### Statistical analysis

Data are presented as mean ± SEM. The parameters of interest at baseline were compared using chi-square or ANOVA based on pre-specified comparisons among the groups (surgical vs. controls, and GB vs SG). The effect of administration of sitagliptin during OGTT vs. control studies of OGTT and the group effect (GB, SG, and CN), as well as their interaction on experimental outcomes, were analyzed using repeated measured ANOVA. The relative changes in the outcomes from control to sitagliptin study were compared using ANOVA based on pre-specified comparisons among the groups. Association among parameters were performed using Spearman correlation. Statistical analyses were performed using SPSS 28 (SPSS Inc., Chicago, IL). The STROBE cross sectional reporting guidelines were used(29).

## RESULTS

### Subjects (Table.1)

**Table 1.**
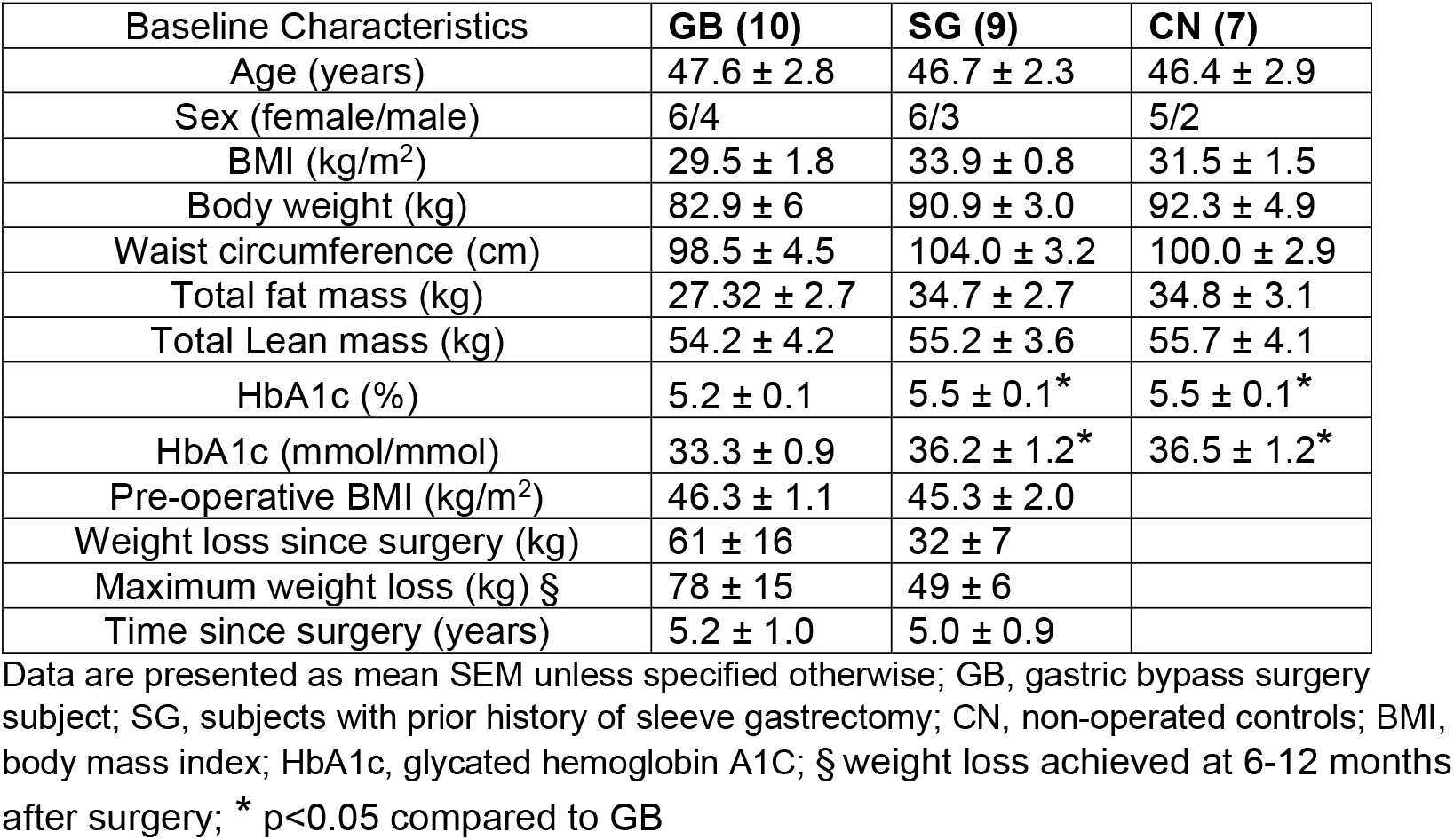
Baseline characteristics of study subjects.

The GB, SG, and healthy controls (CN) had similar BMI, waist circumference, fat/lean body mass. The age and female to male ratio was similar among 3 groups. Among the surgical groups, pre-operative BMI, weight loss and time since GB and SG did not differ significantly. Glycated hemoglobin A1c values ranged from 4.9-5.7% in GB, 4.9-5.9% in SG, and 5.1-6% in CN.

### Glucose and glucose kinetics (Table.2)

**Table 2.** Glucose flux and beta-cell secretory responses to oral glucose ingestion with and without sitagliptin administration in GB, SG and CN subjects.

| Variables | Time (min) | Sitagliptin study |  |  | Control study |  |  | Statistical test |  |  |
| --- | --- | --- | --- | --- | --- | --- | --- | --- | --- | --- |
|  |  | GB | SG | CN | GB | SG | CN | T | G | I |
| Glucose (mmol/L) | Basal | 5.5 ± 0.2 | 5.3 ± 0.1 | 5.5 ± 0.2 | 5.4 ± 0.1 | 5 ± 0.2 | 5.7 ± 0.2 |  |  |  |
|  | Pre-OGTT | 5.1 ± 0.1 | 5 ± 0.2 | 5.2 ± 0.2 | 5.3 ± 0.1 | 4.9 ± 0.2 | 5.4 ± 0.1 | 0.23 | 0.30 | 0.14 |
|  | Nadir | 3.2 ± 0.2 | 4.4 ± 0.3 | 5.1 ± 0.2 | 3.6 ± 0.2 | 4.3 ± 0.3<sup>§</sup> | 5.2 ± 0.2<sup>§</sup> | 0.28 | **0.00** | 0.17 |
|  | Peak | 11.2 ± 0.5 | 9.9 ± 0.4 | 8.2 ± 0.2 | 11.8 ± 0.5 | 10.5 ± 0.3<sup>§</sup> | 9.6 ± 0.3<sup>§</sup> | **0.01** | **0.00** | 0.53 |
| AUC<sub>Glucose</sub> (mmol.min) | (0-60min) | 230 ± 25 | 181 ± 16 | 110 ± 6 | 255 ± 27 | 215 ± 14 | 133 ± 11<sup>§</sup> | **0.02** | **0.00** | 0.93 |
|  | (0-180min) | 162 ± 33 | 331 ± 58 | 297 ± 16 | 226 ± 42 | 409 ± 37<sup>§</sup> | 367 ± 36<sup>§</sup> | **0.01** | **0.00** | 0.97 |
| ISR (pmol.m<sup>-2</sup>.min) | Basal | 167 ± 32 | 132 ± 18 | 112 ± 10 | 157 ± 27 | 122 ± 17 | 104 ± 6 | 0.15 | 0.30 | 0.90 |
|  | Pre-OGTT | 151 ± 29 | 124 ± 12 | 107 ± 10 | 144 ± 29 | 103 ± 12 | 91 ± 7 | **0.00** | 0.30 | 0.50 |
|  | Peak | 3155 ± 447 | 1698 ± 186 | 923 ± 107 | 2555 ± 511 | 1396 ± 191 | 856 ± 92<sup>*</sup> | **0.01** | **0.00** | 0.17 |
| AUC<sub>ISR</sub> (nmol.m<sup>-2</sup>) | (0-60min) | 100.5 ± 14.2 | 55.5 ± 7.6 | 29.2 ± 3.4 | 81 ± 10.3 | 47.8 ± 6.5 | 25.4 ± 3.1<sup>*</sup> | **0.01** | **0.00** | 0.18 |
|  | (0-180min) | 115 ± 12.5 | 101.8 ± 18 | 69.2 ± 8.6 | 91 ± 6.9 | 72.1 ± 7.3 | 55.9 ± 5.3<sup>*</sup> | **0.01** | **0.02** | 0.70 |
| RaO (g) # | (0-60min) | 17.5 ± 1.3 | 15.4 ± 0.9 | 11.5 ± 0.6 | 18.6 ± 1.2 | 17.5 ± 1.3 | 13.6 ± 1<sup>*</sup> | **0.01** | **0.00** | 0.75 |
| RaO (g) # | (0-180min) | 24.5 ± 1.9 | 25 ± 3.2 | 30.2 ± 1.3 | 24.6 ± 1.4 | 27.2 ± 2.7 | 31.7 ± 1.5<sup>§</sup> | 0.20 | 0.08 | 0.66 |
| EGP (g) # | (0-60min) | 4.9 ± 0.7 | 4.7 ± 0.8 | 4.9 ± 0.5 | 4.8 ± 0.6 | 4.6 ± 0.5 | 4.7 ± 0.3 | 0.67 | 0.96 | 0.99 |
| EGP (g) # | (0-180min) | 18.9 ± 1.9 | 11.9 ± 1.3 | 10.3 ± 0.9 | 16.3 ± 1.6 | 12.6 ± 1.3 | 10.6 ± 0.9 | 0.43 | **0.00** | 0.11 |
| Rd (g) # | (0-60min) | 21.3 ± 1.6 | 15.3 ± 1.6 | 12.7 ± 0.5 | 20.9 ± 1.2 | 15.2 ± 1.3<sup>§</sup> | 12.9 ± 1.5<sup>§</sup> | 0.89 | **0.00** | 0.95 |
| Rd (g) # | (0-180min) | 48.8 ± 3.2 | 41.7 ± 4.8 | 42.1 ± 1.6 | 47.6 ± 2 | 45.8 ± 3.8 | 44.4 ± 2.2 | 0.12 | 0.44 | 0.13 |
| HOMA-IR | Basal | 1.6 ± 0.3 | 1.8 ± 0.2 | 2.4 ± 0.4 | 1.6 ± 0.3 | 1.6 ± 0.2 | 2.3 ± 0.2 | 0.88 | 0.09 | 0.76 |
| Insulin sensitivity (L.min<sup>-1</sup>.kg<sup>-1</sup> per µU.L<sup>-1</sup>) | Pre-OGTT | 0.3 ± 0.1 | 0.2 ± 0 | 0.2 ± 0.0 | 0.4 ± 0 | 0.3 ± 0.1 | 0.2 ± 0.0<sup>§</sup> | 0.30 | 0.06 | 0.33 |
|  | (0-180min) | 50 ± 4.9 | 17.9 ± 2.9 | 12.5 ± 2 | 47.7 ± 5.5 | 27.6 ± 4.5<sup>§</sup> | 14 ± 2.4<sup>§</sup> | 0.23 | **0.00** | 0.18 |
| Insulin clearance (L.min<sup>-1</sup> m<sup>-2</sup>) | Basal | 4.2 ± 0.5 | 2.8 ± 0.4 | 1.7 ± 0.1 | 3.9 ± 0.5 | 2.4 ± 0.2 | 1.6 ± 0.1<sup>*</sup> | 0.18 | **0.00** | 0.74 |
|  | (0-180min) | -153 ± 39 | - 67 ± 64 | - 22 ± 19 | - 116 ± 57 | - 42 ± 64 | - 22 ± 16 | 0.30 | 0.80 | 0.48 |

surgical controls; ISR, insulin secretion rate; RaO, systemic appearance of ingested glucose; SGU, splanchnic glucose uptake; EGP, endogenous glucose production; Rd, glucose disappearance; # cumulative values; Statistical effects p values (treatment [control/sitagliptin], group status [GB/SG/CN], and their interaction) are provided in the last 3 columns - T, treatment vs control; G, group status; I, interaction. \* p<0.05 compared to surgical groups; § p<0.05 compared to GB

Fasting plasma glucose was similar among 3 groups (Table.2). As expected, the post-OGTT nadir glucose concentration was lower and glucose excursion (max – min) and AUC_Glucose1h_ were larger in GB compared to SG subjects (p<0.05), and in SG versus non-operated controls (p<0.05; Fig.1a). Despite the higher early rise in plasma glucose after GB, AUC_Glucose3h_ was lower than that in SG and CN (p<0.01; Fig.1a).

**Figure 1.**
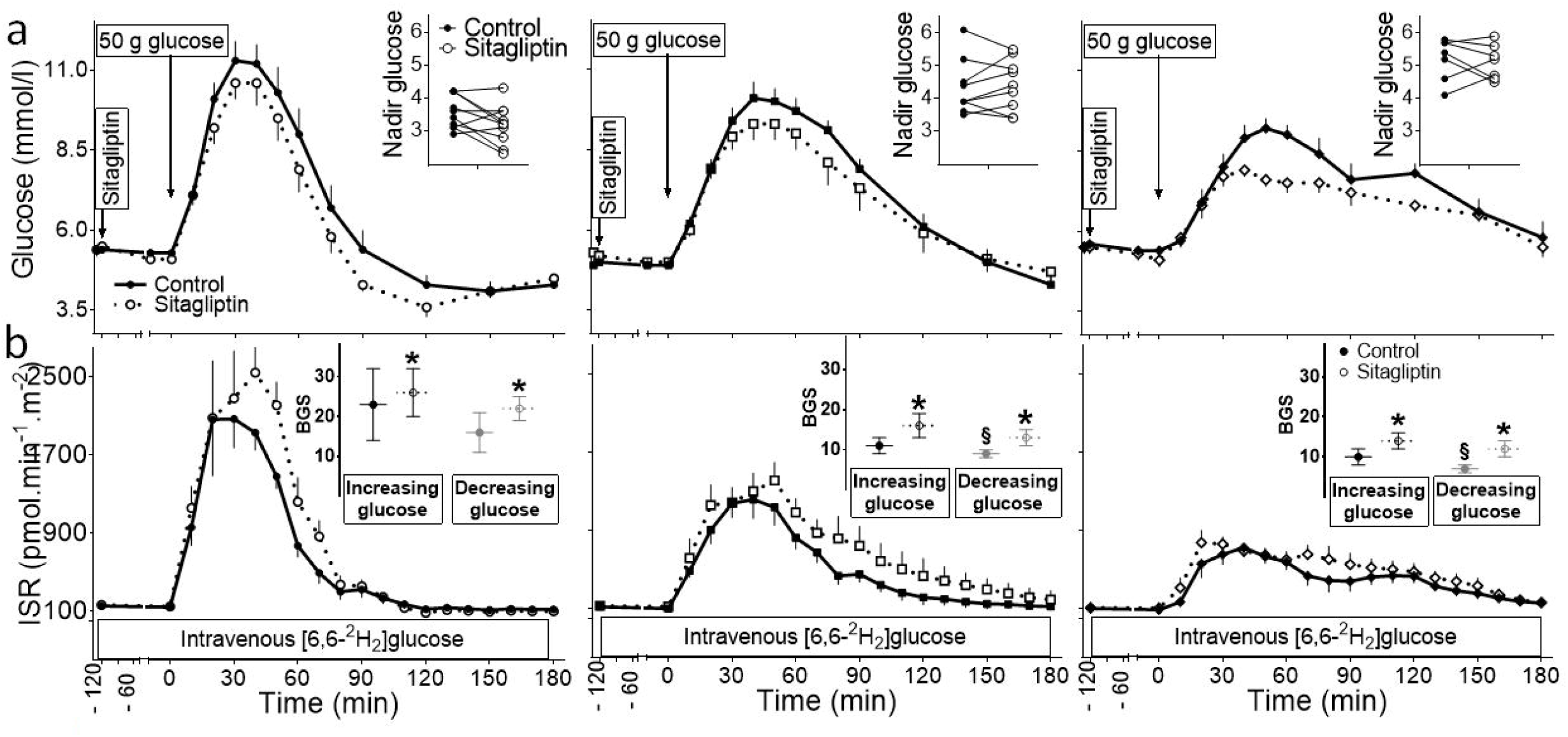
(a) Plasma glucose and (b) insulin secretion responses to oral glucose ingestion with (dashed line) and without (solid line) administration of sitagliptin in subjects who underwent gastric bypass (left panel) or sleeve gastrectomy (middle panel) and non-operated controls (left panel). The corresponding changes in nadir glucose levels as well as β-cell glucose sensitivity during the initial phase of the OGTT and the latter part of the OGTT are shown (insets). *P < .05 compared with control study; § compared with GB.

Sitagliptin lowered post-OGTT glucose response similarly in all 3 groups (p<0.05; Fig.1a) with AUC_Glucose3h_ reduction of 35±38% in GB, 20±12% in SG, and 14±10% in CN. However, nadir glucose levels decreased to a larger extent in GB than SG or CN (relative reduction in nadir glucose of 10±4% in GB versus –3±4% in SG, and 0±5 % in CN; p<0.05). Three subjects in GB, but none in SG or CN, had plasma glucose concentrations between 3.0 and 3.3 mmol/l. Following administration of sitagliptin 5 patients in GB groups had nadir glucose levels <3.3 mmol/l with 3 of the 5 becoming hypoglycemic (glucose<3 mmol/l).

In parallel to glycemia, the early rate of rise in oral glucose appearance (*RaO*) was larger in GB and SG than CN (p<0.05; Fig.2b). There was a trend toward higher *RaO* over the 3-hour period in CN compared to surgical subjects (p=0.08; Fig.2b). DPP4i similarly reduced early *RaO* in all 3 groups (p<0.05) but did not change the total *RaO* (Fig.2b).

**Figure 2.**
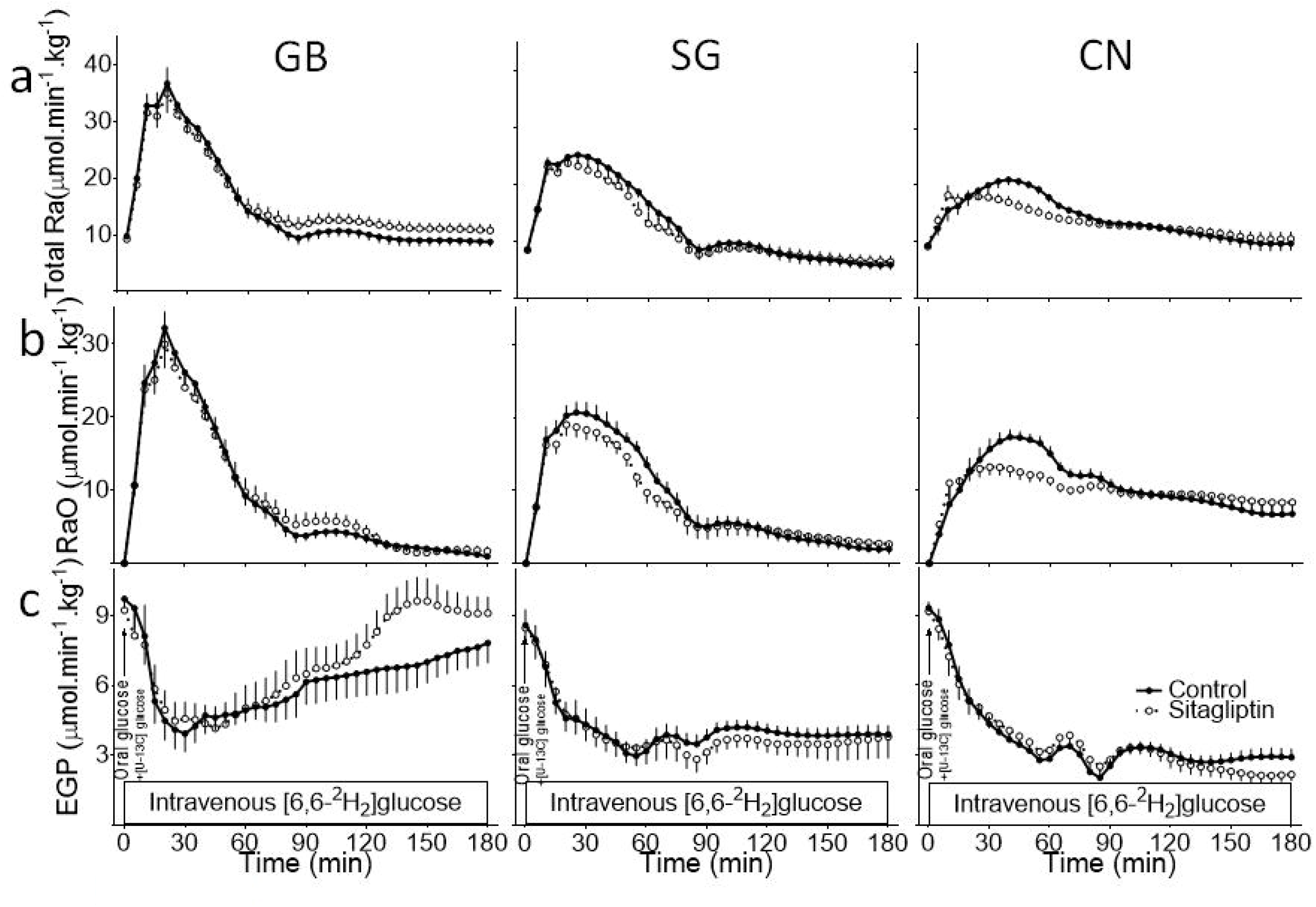
The rates of (a) total glucose appearance (*Ra*), (b) ingested glucose appearance into circulation (*RaO*), and (c) endogenous glucose production (*EGP*) during OGTT with (dashed line) and without (solid line) administration of sitagliptin in subjects who underwent gastric bypass (left panel) or sleeve gastrectomy (middle panel) and non-operated controls (left panel).

Following an overnight fast, steady-state conditions prevail, and the rate of total body glucose utilization (*Rd*) equals the rate of endogenous glucose production (*EGP*), and were similar among GB, SG, and CN (9.7±0.4, 8.6±0.7, and 9.3±0.3 μmol/kg/min, in GB, SG, and CN; Fig.2).

*EGP* similarly declined by 75-80% in response to glucose ingestion in all 3 groups, and this early OGTT-induced *EGP* suppression was not affected by DPP4i. In SG and CN, post-OGTT *EGP* suppression was persistent while in GB subjects, *EGP* rose within 60 minutes of glucose intake, reversing toward premeal levels by 180 minutes (Fig.2c). DPP4i had no significant effect on *EGP* after OGTT in SG and CN but increased post-OGTT *EGP*_*3h*_ in GB subjects (relative change in *EGP*_*3h*_ with and without DPP4i: 19.3±11.5% in GB, –5.8±6.6% in SG, and –0.1±9.5% in CN; p=0.08; Fig.2).

The early rates of glucose disposal (*Rd*_*1h*_) were larger in GB compared to SG and CN (p<0.001), but *Rd*_*3h*_ was similar among the 3 groups. There was no difference in the rates of *Rd* between the studies with and without sitagliptin.

### Incretins and gastric emptying

While fasting level of total GLP-1 (*t*GLP-1) was similar among 3 groups, prandial concentrations were much larger in GB versus SG or CN (p<0.001; Fig.3b). DPP4i administration reduced *t*GLP-1, particularly in GB and CN (p<0.05 for interaction; Fig.3b). Total GIP (*t*GIP) concentrations before and after oral glucose ingestion were similar among 3 groups; and prandial GIP was similarly reduced by DPP4i (p<0.001; Fig.3c).

**Figure 3.**
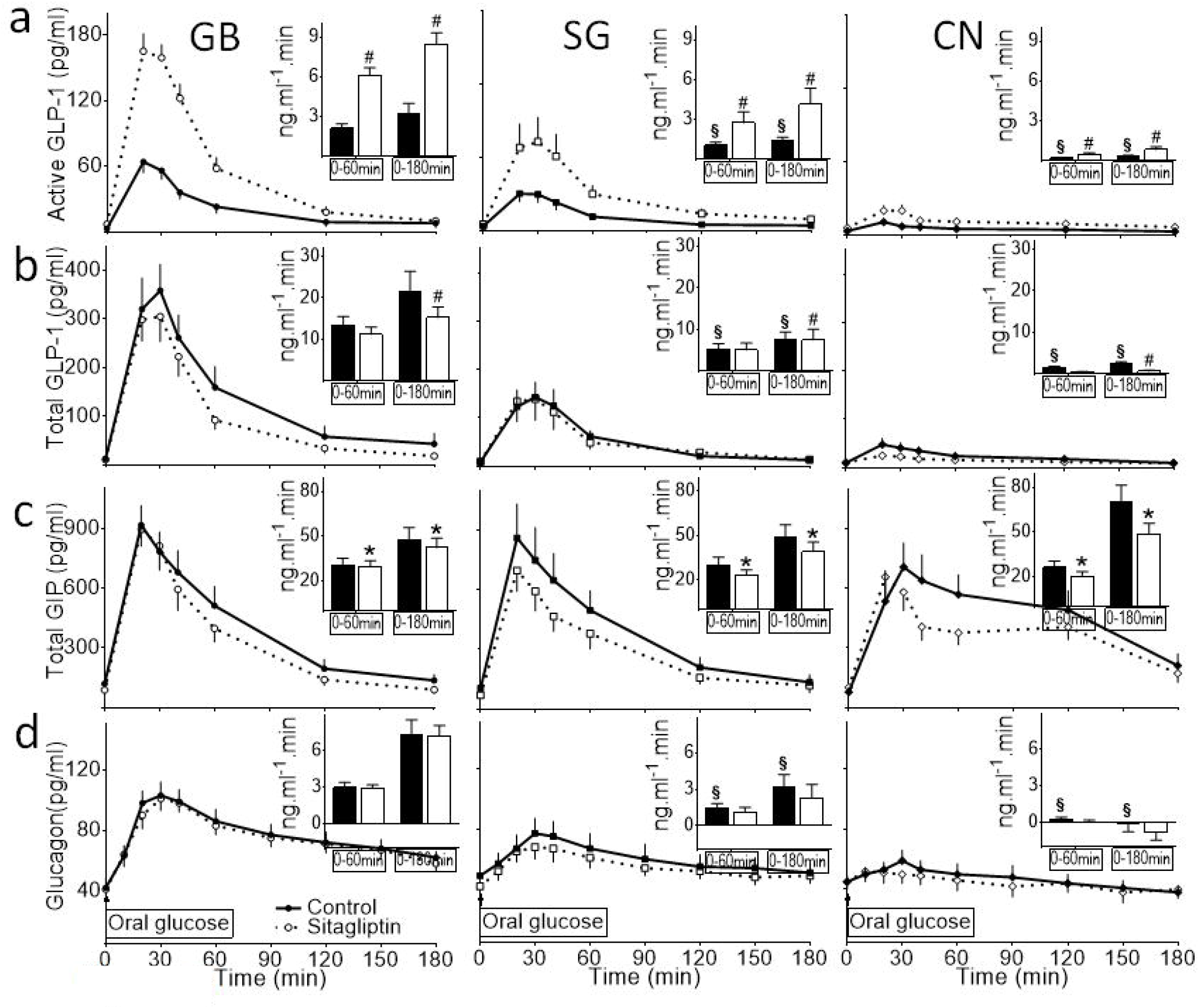
Plasma concentrations of (a) bioactive GLP-1, (b) total GLP-1, (c) total GIP, (d) glucagon during OGTT with (dashed line) and without (solid line) administration of sitagliptin in subjects who underwent gastric bypass (left panel) or sleeve gastrectomy (middle panel) and non-operated controls (left panel). The corresponding AUCs from 0 to 60 min and from 0 to 180 min are shown (insets). * P < 0.05 compared with control study; § P< 0.05 compared with GB; # P < 0.05 for interaction.

Despite similar baseline concentration of *active* GLP-1 (*a*GLP-1) among 3 groups, GB subjects had greater post-OGTT concentrations of *a*GLP-1 than SG, and the response in SG was greater than in CN (p<0.05; Fig.3a). Sitagliptin enhanced the post-OGTT *a*GLP-1 concentrations by 2-3 fold in all 3 groups (p<0.001; Fig.3a).

Time to peak plasma acetaminophen concentration was shorter in GB than SG, and in SG versus CN (21±1, 45±9, and 122±12 min in GB, SG, and CN; p<0.001; Supplementary Figure) and C_max_ was larger in surgical than controls (79±9, 57±9, and 41±3 μmol/L in GB, SG, and CN; p<0.001; Supplementary Figure). DPP4i did not influence the time to peak or the C_max_.

### Beta-cell function (Table.2)

Fasting ISR was similar among the 3 groups. Subjects with GB had an earlier and more robust insulin secretory response to oral glucose ingestion compared to SG and CN subjects (p<0.05; Fig.1b). During control studies, the AUC_ISR3h_ was similar in the two surgical groups but was larger in GB than CN subjects (p<0.05). Beta-cell glucose sensitivity during the first part of the OGTT tended to be higher in GB than SG and CN (Fig.1b). The glucose stimulated ISR as glycemia declined in the latter part of the OGTT was significantly greater in GB compared to SG and CN (p<0.05, Fig.1b). Administration of sitagliptin increased premeal ISR as well as the glucose-induced stimulation of β-cell secretion during the early glycemic rise in the first part and during glycemic decline in the latter part of OGTT (p<0.001; Fig.1b).

Beta-cell responsiveness to *a*GLP-1, estimated by the average slope of each subject’s plot of ISR versus *a*GLP-1 concentration, was significantly smaller in GB or SG versus CN during control study (p<0.01; Fig.4a). Administration of sitagliptin reduced the slope of ISR/*a*GLP-1 in all 3 groups (p<0.01 compared to control condition; Fig.4a).

**Figure 4.**
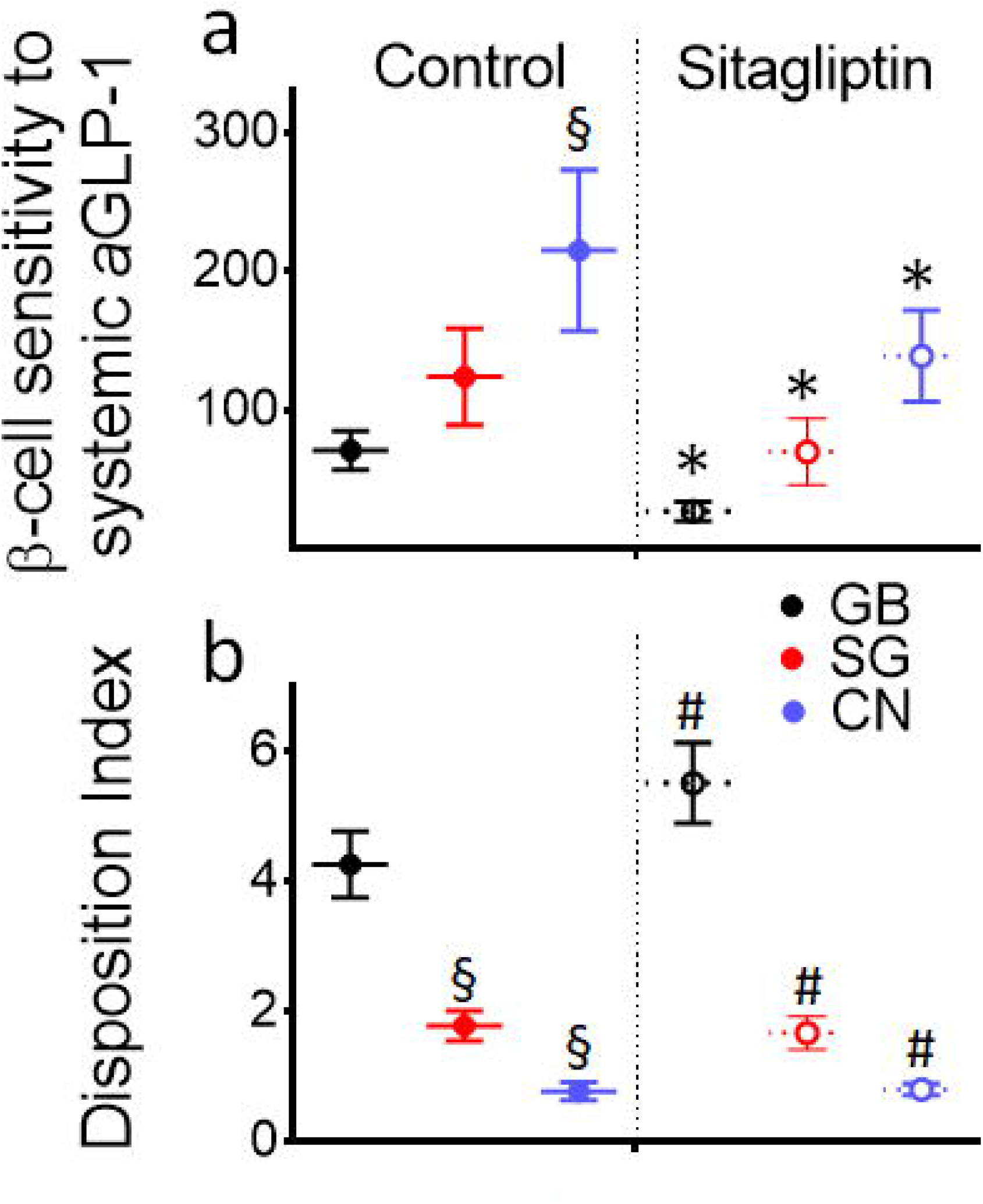
(a) The slope of post-OGTT ISR plotted against increasing plasma concentrations of active GLP-1 (*a*GLP-1) (b) disposition index during control (solid line) and sitagliptin (dashed line) conditions in subjects who underwent gastric bypass (black line) or sleeve gastrectomy (red line) and non-operated controls (blue line). * P < 0.05 compared with control study; § P<0.05 compared with GB; # P < 0.05 for interaction.

Disposition index (DI) was significantly larger in GB versus SG, and in SG versus CN during control studies (p<0.05; Fig.4b). Inhibition of DPP4 increased DI in GB without any effect in SG or CN (P<0.05 interaction; Fig.4b).

### Insulin sensitivity and clearance (Table.2)

Basal and pre-OGTT insulin sensitivity were not significantly different among the groups; post-OGTT insulin sensitivity was larger in GB compared to SG or CN but remained unaffected by DPP4i administration. While baseline insulin clearance was greater in GB compared to SG and CN (p<0.05), there was no significant differences in prandial insulin clearance among the groups. Administration of sitagliptin did not change pre- or post-OGTT insulin clearance.

### Glucagon

Fasting glucagon was similar among the 3 groups. Post-OGTT glucagon concentrations were larger in GB compared to SG and in SG versus non-operated controls (Fig.3d). Glucagon levels before and after OGTT were unaffected by DPP4i.

### Heart rate

Heart rates were similar in fasting and post-OGTT period in 3 groups. Sitagliptin enhanced HR after oral glucose ingestion (average post-OGTT heart rates during control versus sitagliptin study: 72±3, 71±3, and 69±3 bpm in GB, SG, and CN versus 76±2, 76±3, and 73±3 bpm; p<0.01)

### Relationship between post-OGTT DI and nadir glucose with glucose kinetics and hormonal response

Changes in nadir glucose caused by DPP4i was inversely associated with the corresponding changes in DI (r= –0.7; p<0.001) and *EGP* (r= –0.6; p<0.001) in each subject. The magnitude of change in DI and in nadir glucose caused by DPP4i was not related to the changes in plasma levels of *t*GLP-1, *t*GIP, or *a*GLP-1 or baseline characteristics such as age, BMI, or lean mass. Further, post-OGTT *EGP* levels were inversely associated with corresponding nadir glucose concentrations during control (r = –0.5; p<0.05) and sitagliptin studies (r = –0.6; p < 0.001).

## DISCUSSION

The present study was designed to determine the extent to which bariatric surgeries (GB and SG) alter the effect of enhanced plasma concentrations of endogenous incretins by sitagliptin administration on β-cell secretory response to oral glucose ingestion, as well as incretin sensitivity of beta-cells, and to evaluate the differences between GB and SG on this outcome. Consistent with our previous studies with exogenous incretin infusion, we found that β-cell sensitivity to increasing plasma concentrations of *active* GLP-1 after oral glucose ingestion is smaller in GB subjects than in SG and in SG versus CN, despite an opposite hierarchy in β-cell responsiveness to glucose or disposition index across the groups (Fig.4, left panels). Increasing plasma concentrations of *a*GLP-1 by 2-3 fold with DPP4i diminished the ISR/*a*GLP-1 slope even further, but increased β-cell secretory response (disposition index) only in GB and not SG or CN (Fig.4, right panels). As a result, glycemic reducing effect of DPP4i in the latter part of the OGTT (60-180 min) was predominantly observed in GB (Fig.1a). These findings indicate a differential role for the rerouted gut after GB versus SG on prandial glycemic and β-cell effects of *endogenous* incretins in non-diabetic individuals and challenge the traditional model of endocrine action of GLP-1 in humans.

In non-diabetic healthy controls without prior history of GI surgery, prandial glycemic reducing effect of acute inhibition of DPP4 has previously been documented(30, 31, 32, 33). In these studies, glycemic effect of DPP4i is partly mediated by increased β-cell response(30, 31, 32, 33). Although, contradictory to these findings, a study that measured glycemic and insulin secretory response to oral and IV glucose administration with and without sitagliptin in non-diabetic individuals found no effect of DPP4i on glucose or ISR in fasting or fed states(34).

While the reason for this inconsistency in β-cell effects of DPP4i in previous research is unclear, in the current study, administration of sitagliptin significantly reduced the post-OGTT glycemia by increasing β-cell secretion in non-diabetic subjects with and without prior history of bariatric surgery. More specifically, inhibiting DPP4 in this study promoted both fasting and postprandial insulin secretory response to the early glycemia rise (β-cell glucose sensitivity) as well as to the glycemic decline during the second phase of glucose absorption (Fig.1b). Further, GB subjects had the most prominent glycemic effect of DPP4i in the latter part of glucose absorption during sitagliptin study. Also, post-OGTT disposition index was increased by inhibition of DPP4 activity in GB and not in SG or CN (Fig.4b).

The observation that augmented incretin levels enhance insulin secretion independent of glucose levels is aligned with previous studies in that glycemic reducing effect of DPP4i was observed during protein or fat ingestion where glucose concentrations dipped below the baseline values in non-diabetic subjects without GI surgery(31). Similarly, in another study, 4-weeks of DPP4i administration compared to placebo in patients with T2D resulted in larger endogenous insulin secretion during insulin-induced hypoglycemic clamp (5 and 2.5 mM)(35). These findings challenge the traditional model of glucose-dependency of incretins in the prandial state. In fact, the assumption of muted insulinotropic effects of incretins at normal glucose levels is mainly derived from studies of exogenous GIP(36) or GLP-1 infusion(37), which involved increasing systemic levels, not portal incretin concentrations.

The rate of nutrient delivery to the gut plays a major role in regulation of prandial incretin(38). Consistent with prior reports(39), in our study, GIP concentrations rose more rapidly after oral glucose ingestion in GB and SG compared to CN, but the overall GIP response did not differ among the groups. As expected(24, 40), post-OGTT GLP-1 response was larger in GB than SG, and in SG compared to CN. A 2-3-fold increase in plasma concentrations of *a*GLP-1 by DPP4i in our experiment is similar to what previously has been reported in patients with GB(41) or non-operated control(31, 33), suggesting that DPP4 activity is inhibited as planned. Sitagliptin, also, reduced *t*GLP-1 and *t*GIP levels likely due to negative feedback previously described after glucose ingestion(22).

Aligned with prior reports(30, 41), we did not find any association between relative changes in β-cell response induced by DPP4i and those of plasma *a*GLP-1 or *t*GLP-1 concentrations. During control study, β-cell sensitivity to increasing systemic levels of *a*GLP-1 during the first phase of OGTT was 2-3 times smaller in GB and SG than controls despite a larger post-OGTT glycemic rise in surgical subjects. This observation is consistent with our previous report that β-cell responsiveness to *exogenous* GLP-1 administration during fixed hyperglycemia was significantly reduced after GB(23). The assumption here is that the β-cells respond to massive elevation in prandial GLP-1 secretion after GB, and to lesser magnitude after SG, protecting against hypoglycemia. However, increasing plasma concentration of *a*GLP-1, in the current study, reduced *a*GLP-1 sensitivity of β-cells even further (Fig.4A), and yet DI was increased reducing nadir glucose concentration in GB-treated subjects (Fig.4B). Therefore, blunted sensitivity to GLP-1 in presence of elevated plasma GLP-1 concentrations appears to protect against hypoglycemia in SG and CN but not in GB subjects, who remain susceptible to a substantial insulinotropic effect of GLP-1,

These findings also raise the question as to what extent the insulinotropic effects of GLP-1 (during control and sitagliptin studies) is due to hormonal action of intestinally secreted peptide, which makes up majority of circulatory *t*GLP-1 and subsequently *a*GLP-1. The most robust glycemic and β-cell effects of endogenous GLP-1 in our experiment, particularly in GB subjects, were observed during post-OGTT nadir glycemic levels where the plasma concentrations of GLP-1 return to baseline. This is consistent with a recent study which reported a significant increase in nadir glucose concentrations, as well as AUC glucose after the first hour of meal study, after GLP-1R blockade in GB and SG(19).

DPP4 has a widespread organ distribution, including pancreatic α-cells(42). Inhibition of islet DPP4 activity in murine and human isolated islets has shown to increase insulin secretion mediated by GLP-1(42). Furthermore, in a rodent model of sleeve gastrectomy, the pancreatic rather than intestinally secreted GLP-1 was the mediator of glycemic beneficial effects of surgery(43). Therefore, it is plausible that after bariatric surgery, β-cell effects of DPP4i, especially during hypo- or euglycemic conditions in the fed state, are mediated by the paracrine action of GLP-1 produced in the pancreas rather than intestinally secreted peptide. Additionally, the visceral afferent neurons, mainly vagal, in the portal artery express the GLP-1R and play a role in the regulation of glucose metabolism(44). It remains to be investigated as to what extent paracrine or vagally mediated GLP-1 actions plays a role in enhanced prandial β-cell secretion after GB or SG.

Beyond these important differences in β-cell function, GB subjects had the smallest post-OGTT *EGP* suppression, especially from 60-180 minutes, compared to SG and CN despite a larger insulin secretory response. In response to sitagliptin administration there was a lower OGTT-induced *EGP* suppression (90-180 min) in GB compared to the control condition, while prandial *EGP* remain suppressed in SG and CN (Fig.2c). This biphasic pattern of prandial *EGP* after GB has previously been reported by others who compared prandial *EGP* in GB versus SG(40), in GB versus non-operated controls(20), and before and after GB(1). It’s largely unknown whether prandial hyperglucagonemia in GB subjects during the second part of OGTT plays a role in the *EGP* rise. However, in this study, the *EGP* rise in GB was much greater after sitagliptin despite an increased post-OGTT insulin secretion without any change in plasma glucagon concentration, suggesting that factors beyond prandial hormonal response are involved.

There are limitations to this study that warrant consideration. The cross-sectional design rather than longitudinal study imposes limitation in within-subject comparison of the effect of bariatric surgery on the outcomes. However, our method allowed us to examine the effect of bariatric surgery beyond the first 2 years, when our subjects were completely adapted to metabolic effect of surgery with maximal weight loss achieved and maintained at the time of the experiment. A previous study has found no glycemic effect for 200 mg sitagliptin administration in non-diabetic GB subjects within 5 months of surgery by comparing post-meal glucose and insulin secretion(41). The discrepancy in the findings suggests that metabolic adaptation of bariatric surgery on gut function or pancreatic β-cell response take longer than the first few months after these procedures. Our participants were enrolled consecutively based on inclusion criteria. Moreover, we did not measure *active* GIP, however, it can be assumed that *a*GIP is also increased in parallel to *a*GLP-1. However, it is unlikely that increased disposition index as observed in in GB subjects in the present study is mediated by enhanced *a*GIP since blocking GIPR in patients with GB has no effect on post-meal ISR or glucose concentrations(19). Furthermore, our findings can’t give insights into glycemic effects of DPP4i in bariatric subjects with diabetes since our subjects had no known history of diabetes.

In summary, our findings indicate that inhibition of DPP4 activity in glucose tolerant subjects: (1) augments the β-cell insulin secretory response to the oral glucose challenge, but more robustly in those with history of GB than SG or non-operated controls; (2) lowers nadir glucose in GB leading to hypoglycemia in 3 of 10 subject but has no effect to reduce nadir glucose in SG or CN; (3) impairs post-OGTT *EGP* suppression in GB subjects during the latter part of glucose absorption. Thus, increasing the plasma incretin concentration by administration of a DPP4 inhibitor shifts the glycemic and insulin profile in asymptomatic GB subjects to mimic the profile in those with post-GB hypoglycemia. Most importantly, and furthermore, consistent with our previous findings, the β-cell sensitivity to increasing concentrations of endogenous GLP-1 is smaller after bariatric surgery, particularly after GB, compared to non-operated controls, indicating a long-term adaptation of gut-pancreas axis after these procedures, likely to protect against hypoglycemia. Finally, the opposite effect of DPP4i on β-cell sensitivity to plasma concentrations of *a*GLP-1 versus disposition index across the groups suggest that insulinotropic effects of GLP-1 after gastric bypass surgery are likely mediated by both hormonal and non-hormonal mechanisms.

## Data Availability

All data produced in the present work are contained in the manuscript.

## Abbreviations

(AUC): Areas under the curve
(BMI): body mass index
(CN): non-operated control subjects
(DPP4i): dipeptidyl peptidase-4 inhibitor
(DI): disposition index
(EGP): endogenous glucose production
(GB): Roux-en Y gastric bypass
(GI): gastrointestinal
(GIP): glucose- dependent insulinotropic peptide
(tGIP): total GIP
(GIR): glucose infusion rate
(GLP-1): glucagon like-peptide 1
(GLP-1R): GLP-1 receptor
(aGLP-1): active GLP-1
(*t*GLP-1): total GLP-1
(HbA1c): glycated hemoglobin A1C
(IC): insulin clearance
(ISR): insulin secretion rate
(*MCR*_*Glucose*_): metabolic clearance rate of glucose
(PP): pancreatic polypeptide
(RaO): systemic appearance of ingested glucose
(Rd): total glucose disappearance
(SG): sleeve gastrectomy
(TRa): total glucose appearance

## ACKNOWLEDGEMENT

We thank Andrea Hansis-Diarte, Nancy Yegge, and John Adams from the Department of Medicine of University of Texas Health at San Antonio for their technical support and nursing staff as well as nutritionist from Bartter Research Unit, Audie Murphy Hospital, STVHCS, for their expert technical assistance. We owe a great debt to our research participants.

## AUTHOR CONTRIBUTION

MS designed and supervised the study, obtained the data, analyzed, and interpreted the data, and wrote the manuscript and takes full responsibility for the work as a whole (guarantor); AG analyzed the data; RP, DT, and SP contributed to conducting studies and obtaining data, RD and AG contributed to interpretation of data and review/editing of the manuscript.

## DISCLAIMER

Parts of this study were presented at American Diabetes Association, 81^st^ Scientific Sessions (virtual). All data generated or analyzed during this study are included in this published article (and its supplementary information files). Authors have nothing to disclose conflicting with the content of current manuscript.

## FIGURE LEGENDS

**Supplementary Figure.**
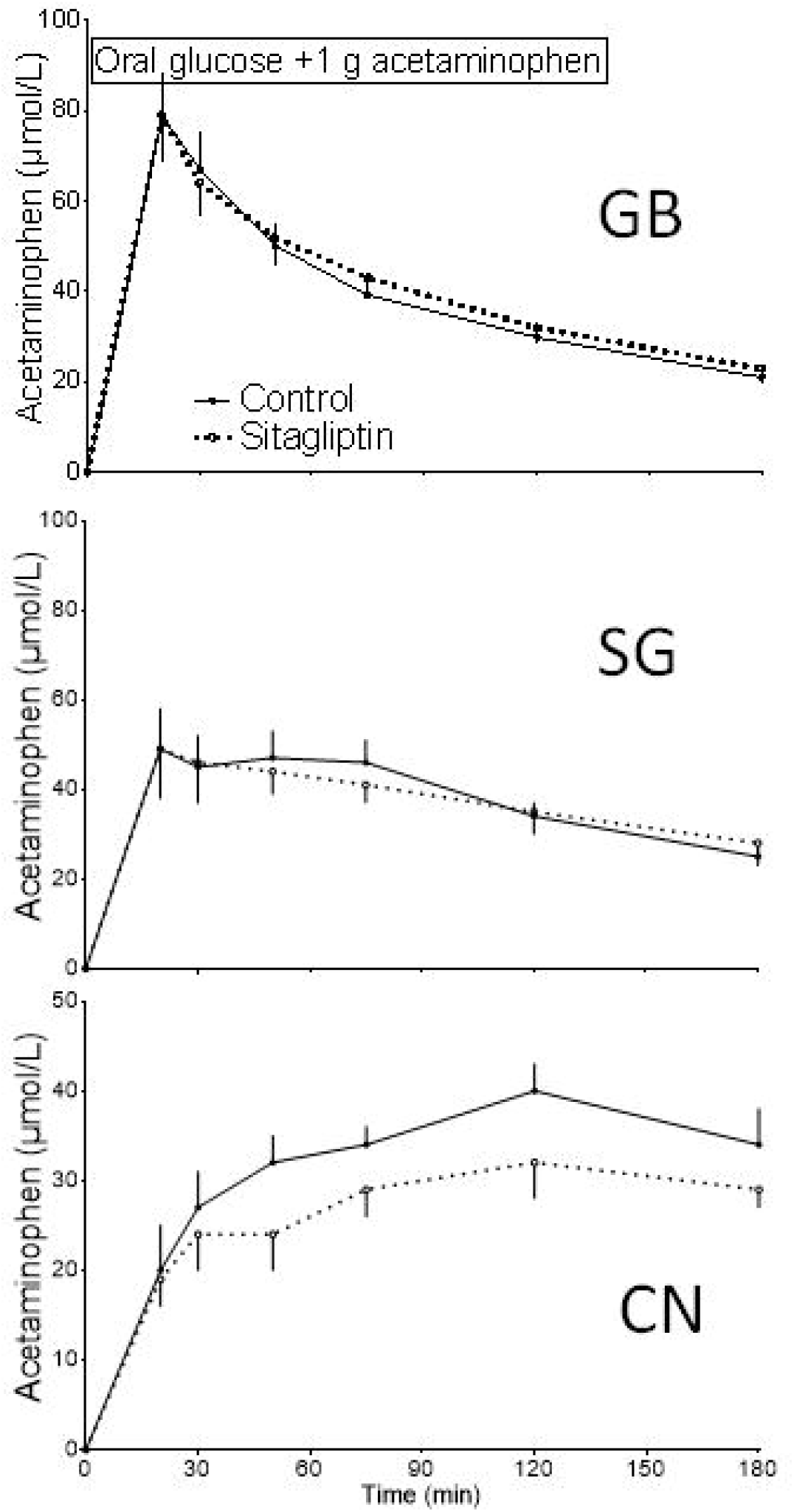
Plasma concentrations of acetaminophen after oral glucose ingestion during OGTT with (dashed line) and without (solid line) administration of sitagliptin in subjects who underwent gastric bypass (top panel) or sleeve gastrectomy (middle panel) and non-operated controls (bottom panel)

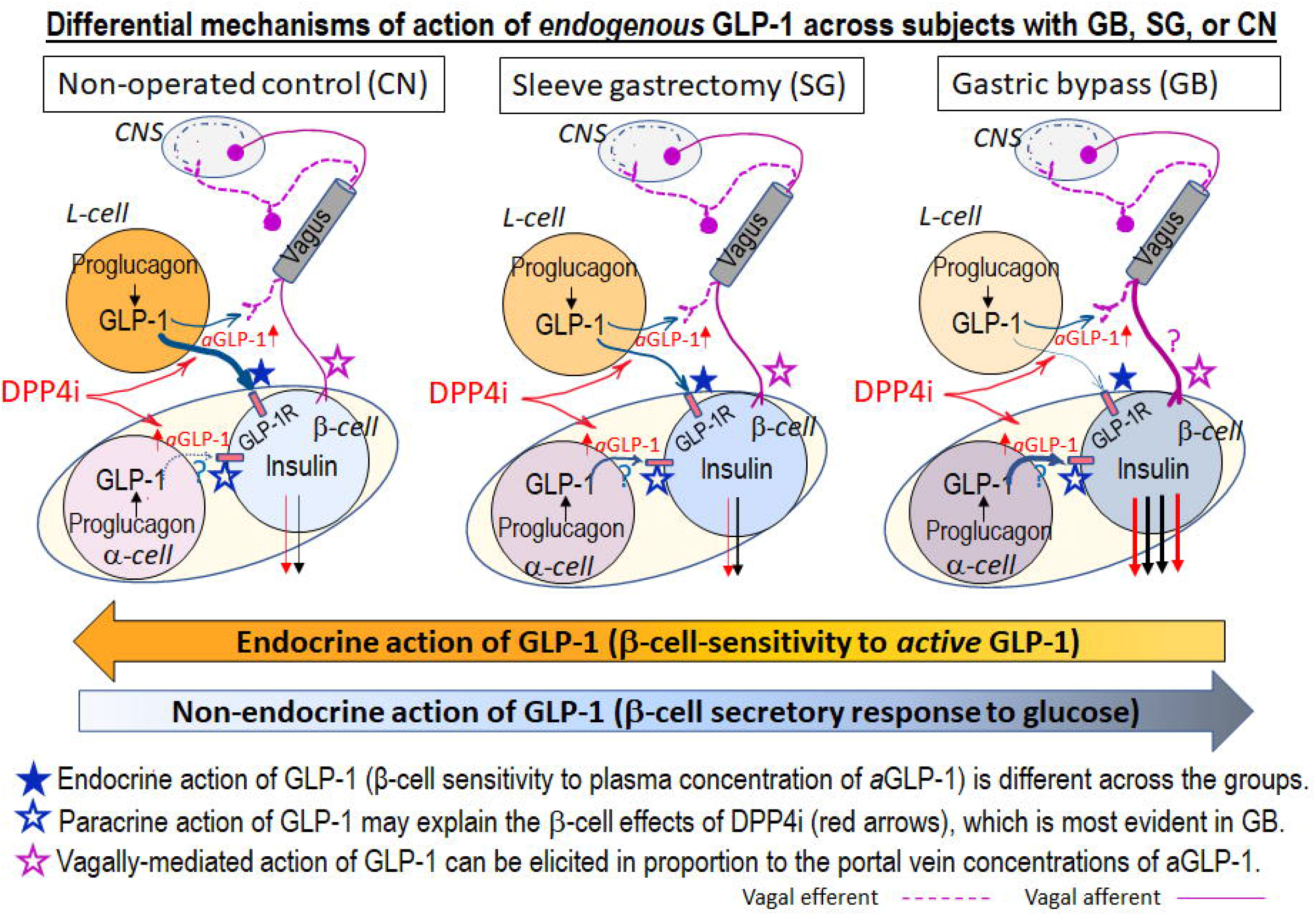

